# Worrisome Emergence of Pan-INSTI Resistance: A Systematic Scoping Review of Dolutegravir Resistance in INSTI-Naïve Patients Post-Therapy Failure

**DOI:** 10.1101/2025.02.01.25320414

**Authors:** Sumit Arora, Nishant Raman, Anirudh Anilkumar, Kuldeep Ashta, N Kisenjang, Charu Mohan

## Abstract

**Introduction:** Dolutegravir (DTG), a second-generation integrase strand transfer inhibitor (INSTI), is widely used in HIV treatment, especially in low-resource settings. Despite its proven efficacy, concerns about DTG resistance mutations (DRMs) have emerged in patients failing dual NRTI + DTG-based ART regimens. This review examines the patterns and frequencies of these DRMs in INSTI-naïve patients.

**Methods:** A systematic scoping review was conducted, synthesizing data from 21 studies (2013–2024) involving 59 INSTI-naïve persons living with HIV-1 (PLH) who experienced virological failure (VF) on dual NRTI + DTG-based ART. Data extraction was undertaken by two independent reviewers, and key information included ART history, DRM profiles, duration of DTG-based ART, and viral load at failure. A qualitative synthesis identified common resistance patterns, geographic distributions, and HIV subtype correlations.

**Results:** The most prevalent DRMs were G118R (42.4%) and R263K (38.9%). G118R, when combined with T66I and E138K, was associated with high-level resistance and pan-INSTI resistance. R263K, frequently occurring alone or with minor mutations, also conferred modest resistance. Resistance patterns varied by HIV subtype, with non-B subtypes showing higher frequencies of G118R and Q148HRK mutations, while R263K predominated in subtype B.

**Conclusion:** Emerging DTG resistance in INSTI-naïve patients, particularly in resource-limited settings, is a cause for concern. G118R and R263K were the most prevalent mutations, with the former leading to pan-INSTI resistance. These findings stress the importance of monitoring resistance patterns, especially in non-HIV-B subtypes, to optimize ART strategies.

## Introduction

Dolutegravir (DTG), a second-generation integrase strand-transfer inhibitor (INSTI), is widely preferred in first- and second-line antiretroviral therapy (ART) due to its superior virological efficacy, favourable safety profile, and reduced pill burden.^[1]^ Recognizing its advantages, the World Health Organization (WHO) recommended DTG-based regimens as the preferred option for first- and second-line ART across all populations in 2019.^[2]^

DTG offers significant advantages over earlier INSTIs. First-generation INSTIs, such as Raltegravir (RAL) and Elvitegravir (EVG), have been in use for many years but are characterized by a modest barrier to resistance. These agents are linked to the rapid development of resistance through single mutations or primary mutations combined with secondary mutations in the integrase enzyme. ^[3–5]^ In contrast, second-generation INSTIs—including Bictegravir (BIC), Cabotegravir (CAB), and particularly DTG—possess a high genetic barrier to resistance and exhibit very low cross-resistance with first-generation INSTIs. ^[5,6]^ This enhanced resistance profile has solidified DTG’s position as a robust option in both first- and second-line ART regimens.

The widespread use of DTG in resource-limited settings—where ART regimens are highly standardized with limited flexibility for optimized background therapy—raises concerns about the potential emergence of resistance. In these settings, the recycling of drugs, restricted access to adherence support, and limited availability of viral-load and resistance testing can exacerbate the problem.^[7]^ Emergence of DTG-selected drug resistance mutations (DRMs) has been reported in up to 3.8% of ART-experienced, INSTI-naïve people living with HIV (PLH) who failed first- or second-line dual NRTI + DTG-based ART regimens in clinical trials, with a median prevalence of 1.5%, and 0.7% in ART naïve PLH receiving first-line dual NRTI+ DTG.^[8]^ While initially considered a rare phenomenon, the development of DRMs following DTG-based ART failure has grown into a significant concern. Resistance to DTG is now being reported at unexpectedly high rates in real-world settings, exceeding those observed in clinical trials.^[8–10]^

This scoping review synthesizes all available studies reporting the emergence of INSTI-associated DRMs in INSTI-naïve PLH experiencing virological failure (VF) while on the WHO-recommended, and widely used dual NRTI + DTG-based first- or second-line ART regimens. By analysing these data, it provides a real-world perspective on the patterns and relative frequencies of DRMs under DTG selective pressure, including mutations associated with pan-INSTI resistance.

## Methods

### Review design

A systematic scoping review was conducted to evaluate emergent pan-INSTI drug resistance mutations (DRMs) in HIV-1 patients receiving dual nucleoside reverse transcriptase inhibitors (NRTI) + dolutegravir (DTG)-based ART, considering diverse clinical and demographic contexts. The aim of the study was to identify and analyse studies reporting on ART-naïve or treatment-experienced, INSTI-naïve persons living with HIV (PLH), and without exposure to third-line ART agents such as Darunavir, or fusion, or entry inhibitors, who developed emergent INSTI drug-resistance mutations (DRMs) after failing dual NRTI+DTG-based ART, with the goal of identifying common patterns of emergent INSTI DRMs in real-world clinical settings.

The review included clinical trials, observational studies, case reports, and grey literature published from 2013 to 2024, aligning with the period of widespread DTG use following WHO recommendations.^[11]^ Data were sourced from PubMed and Google Scholar, with the final search conducted on November 4, 2024. Detailed search strings are provided in the supplementary protocol file.

### Data extraction

Data-extraction was independently performed by two-reviewers using a standardized form, following PRISMA guidelines for scoping reviews.^[12,13]^ Any discrepancies were resolved through discussion, with a third reviewer consulted when necessary. The data were organized in a way that allowed for comparisons across various study types and geographic regions.

### Inclusion Criteria

The review targeted studies reporting on ART-naïve or treatment-experienced but INSTI-naïve individuals who developed emergent INSTI DRMs after virological failure (VF) of dual NRTI + DTG-based ART. Studies were included if they reported full sequences or identified DRMs, ART drug exposure history, duration of DTG-based ART, viral load (VL) at failure, author-defined VF criteria, and DRT methodology. Only DRMs conferring at least low-level resistance (LLR) to INSTIs per Stanford HIVDB version 9.8 were included.^[14]^

For PLH with more than one isolate containing INSTI-associated DRMs, a cumulative count from all isolates was considered, provided the isolates were from the same period of virological failure. Mutations present as part of a mixture with wildtype were also considered mutant. When multiple publications reported HIV-1 isolates with INSTI DRMs from the same clinical trial or cohort, isolates were linked to only one publication, with preference for the primary analysis.

### Exclusion Criteria

The review excluded articles that were not primary research studies, such as reviews, meta-analyses, or studies lacking detailed data on drug resistance mutations or ART history, and those that describe patients infected with HIV-2. Studies that did not provide specific information on the ART regimen used, or failed to report viral load data, were also excluded. In cases where essential data were missing, attempts were made to contact the corresponding authors for clarification or additional information before their exclusion. Grey literature was excluded.

Studies using Next-Generation Sequencing (NGS) or Ultra-deep sequencing were excluded, and for studies reporting both Sanger sequencing and NGS from the same patient, Sanger sequencing results were used to provide a real-world perspective.

### Data Synthesis and analysis

Data synthesis followed the CoCoPop framework. The population included HIV-1 patients on dual NRTI + DTG-based ART, with demographic and clinical details such as geographic region, age, gender, and VL. Conditions included emergent pan-INSTI resistance mutations and VF outcomes. Context covered ART regimens, prior exposure to other INSTIs, and demographic data, with adherence noted where available. A qualitative synthesis identified common resistance patterns, geographic distributions, and HIV subtype correlations.

### Data Handling

The data handling and analysis were carried out using Rayyan for systematic review management, Microsoft Excel for data organization, and SPSS 23 for advanced statistical analysis. Additionally, the review protocol was registered with the Open Science Framework (OSF) on November 3, 2024, ensuring transparency and reproducibility of the review process. ^[15]^

## Results

### Search results

A total of 419 publications were retrieved (293 from PubMed and 126 from Google Scholar) as shown in **Figure-1**. After removing 15 duplicate publications, titles and abstracts of 404 publications were reviewed. Of these, 122 publications were subjected to full-text screening. Ultimately, 21 publications from 19 studies were included, reporting on INSTI-naïve persons living with HIV (PLH) who developed one or more INSTI drug-resistance mutations (DRMs) conferring at least low-level resistance (LLR) to INSTIs while receiving dual NRTI + DTG. These studies comprised data from 59 individual patients.^[16,17,26–35,18,36,19–25]^

**Figure 1:**
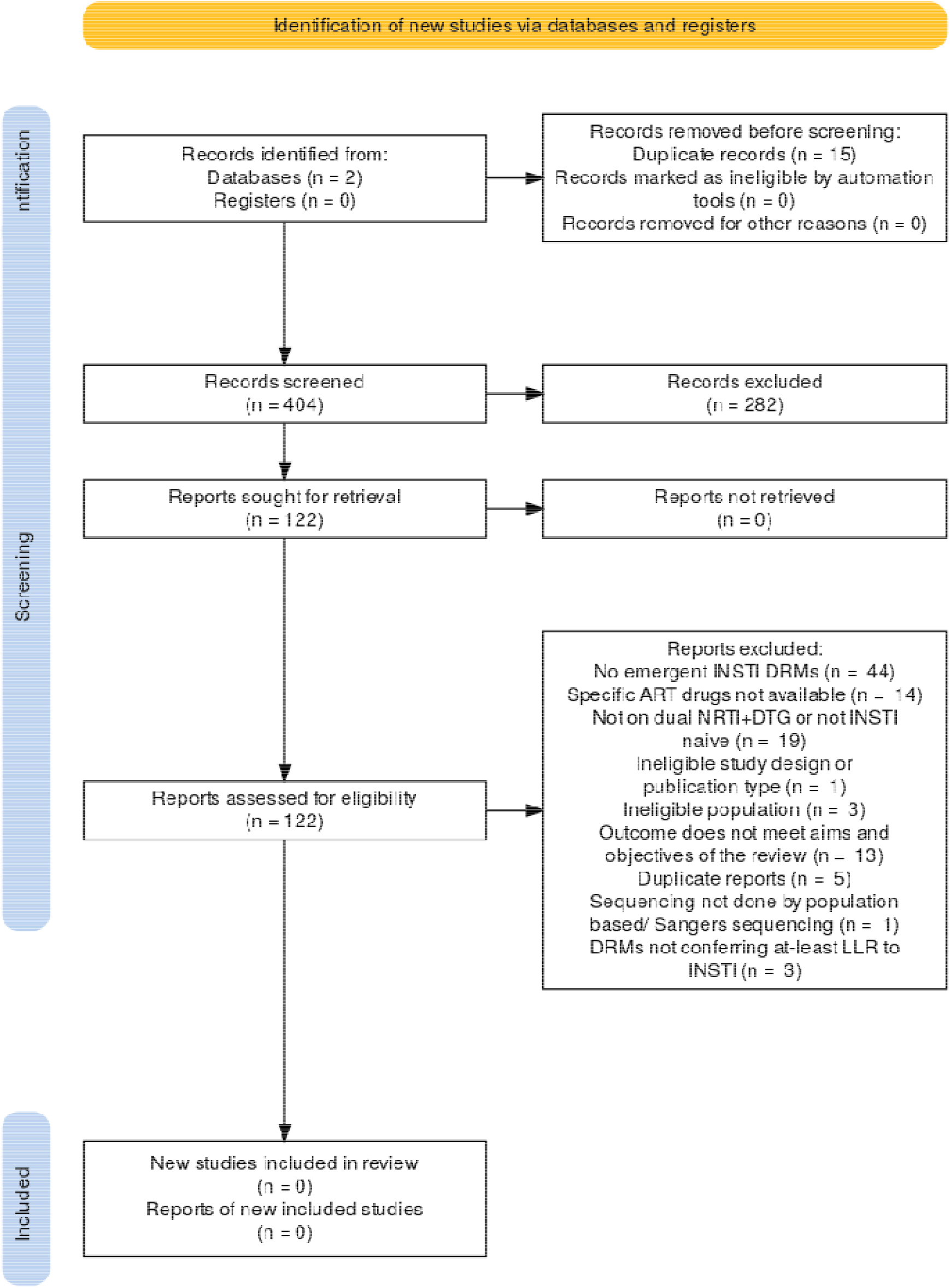
The flow chart summarizes the review process. Exclusions were based on single criteria. Two trials presenting data from the same patients were considered duplicates, while three additional duplicate studies were removed during full-text screening. Fourteen studies were excluded due to the lack of information on current or past ART regimens, despite attempts to contact the authors. **Abbreviations**: ART – Antiretroviral therapy; DRM – Drug-

Among the included studies, five publications reported data on 20 patients in three clinical trials, nine observational studies reported 31 patients, and seven case reports documented eight cases. The geographical distribution includes patients predominantly from African countries (37.3%, n=22), whereas patients from multinational studies involving African, American, Asian and European populations comprised 18.6% (n=11). **Table-1** and **Supplementary Table-S1** summarize the reports of emergent INSTI-DRMs from the included studies.

**Table 1:** Summary of Studies Reporting Emergent INSTI DRMs and Pan-INSTI Resistance in ART-Naïve and INSTI-Naïve PLH on Dual NRTI + DTG Regimens.

| Author | Type of Study | Geographic Area | Regimen During Failure | Past ART Exposure | No. with major INSTI DRMs | No. with pan-INSTI resistance |
| --- | --- | --- | --- | --- | --- | --- |
| Lepik (2017)<sup>[34]</sup> | OBS | Canada | ALD | NAÏVE or EXP but INSTI NAÏVE | 3 | 0 |
| Pena (2018)<sup>[33]</sup> | CR | Spain | TLD | NAÏVE | 1 | 1 |
| Cardoso (2018)<sup>[32]</sup> | CR | Portugal | ALD | PI, NNRTI | 1 | 1 |
| Underwood (2022)<sup>[28,31]</sup> | RCT | Multinational | TED/ TLD | NNRTI | 6 | 6 |
| Mahomed (2020)<sup>[30]a</sup> | CR | South Africa | ALD | PI, NNRTI | 1 | 1 |
| Seatla (2021)<sup>[29]</sup> | OBS | Botswana | ALD | EXP but INSTI NAÏVE | 2 | 2 |
| Paton (2022)<sup>[27,36]</sup> | RCT | Multinational (Africa) | ZLD | NNRTI | 9 | 5 |
| Gil (2022)<sup>[26]b</sup> | OBS | Spain | ALD/ TLD | NAÏVE, NNRTI | 5 | 0 |
| Chirimuta (2022)<sup>[25]</sup> | CR | Zimbabwe | ALD/ TLD | PI, NNRTI | 2 | 2 |
| Kampen (2022)<sup>[24]c</sup> | CR | Netherlands | ALD | PI, NNRTI | 1 | 1 |
| Vavro (2022)<sup>[23]d</sup> | CT | Multinational | ZLD/ TLD/ TED/ TLED | PI, NNRTI | 5 | 5 |
| Bwire (2023)<sup>[22]</sup> | OBS | Tanzania | ALD/ TLD | PI, NNRTI | 3 | 3 |
| Abdulahi (2023)<sup>[21]</sup> | OBS | Nigeria | TLD | EXP but INSTI NAÏVE | 1 | 1 |
| Kamori (2023)<sup>[20]d</sup> | OBS | Tanzania | TLD | NAÏVE, NNRTI | 4 | 3 |
| Farias (2023)<sup>[16]</sup> | CR | Brazil | TLD | NAÏVE | 1 | 0 |
| Malinga(2023)<sup>[35]d,e</sup> | CR | South Africa | ALD | PI | 1 | 1 |
| Armenia (2023)<sup>[19]</sup> | OBS | Multinational (Europe) | ALD/ TLD | PI, NNRTI, NAÏVE | 5 | 1 |
| Diaz (2023)<sup>[18]</sup> | OBS | Brazil | ALD/TLD | NAÏVE | 7 | 2 |
| Tsai (2023)<sup>[17]</sup> | OBS | Taiwan | ALD | NAÏVE | 1 | 0 |

<sup>a</sup> Patient on ALD but took DTG only included. <sup>b</sup> One patient exposed to TLE as failing regimen with recent history of ALD exposure included. <sup>c</sup> Both NGS and Sangers sequencing performed but only results of Sangers sequences interpreted. <sup>d</sup> Studies including paediatric patients. <sup>e</sup> One patient on TLED regimen included. The mutations associated with each patient and other details are available in **Supplementary Table-S1**. Abbreviations: ART – Antiretroviral therapy; DRM – Drug-resistance mutations; EXP – ART experienced; INSTI – Integrase strand transfer inhibitor; PI – Protease inhibitor; NNRTI – Non nucleosidase analogue reverse transcriptase inhibitor; NRTI – nucleosidase analogue reverse transcriptase inhibitor; EXP – ART experienced; CR – Case report; RCT – Randomised controlled trial; CT – Clinical trial; OBS – Observational study. ALD, TED, TLD, TLED, TED represent specific ART regimens.

### Emergent INSTI DRMs

The most commonly detected INSTI DRM in previously INSTI naïve PLH failing dual NRTI + DTG based ART was **G118R**, observed in 42.4% of patients (n=25), followed by **R263K**, which occurred in 38.9% of patients (n=23). **Figure 2** illustrates the relative frequency of signature INSTI DRMs and other major INSTI DRMs across all patients, as well as their contribution to high-level pan-INSTI resistance.

**Figure 2:**
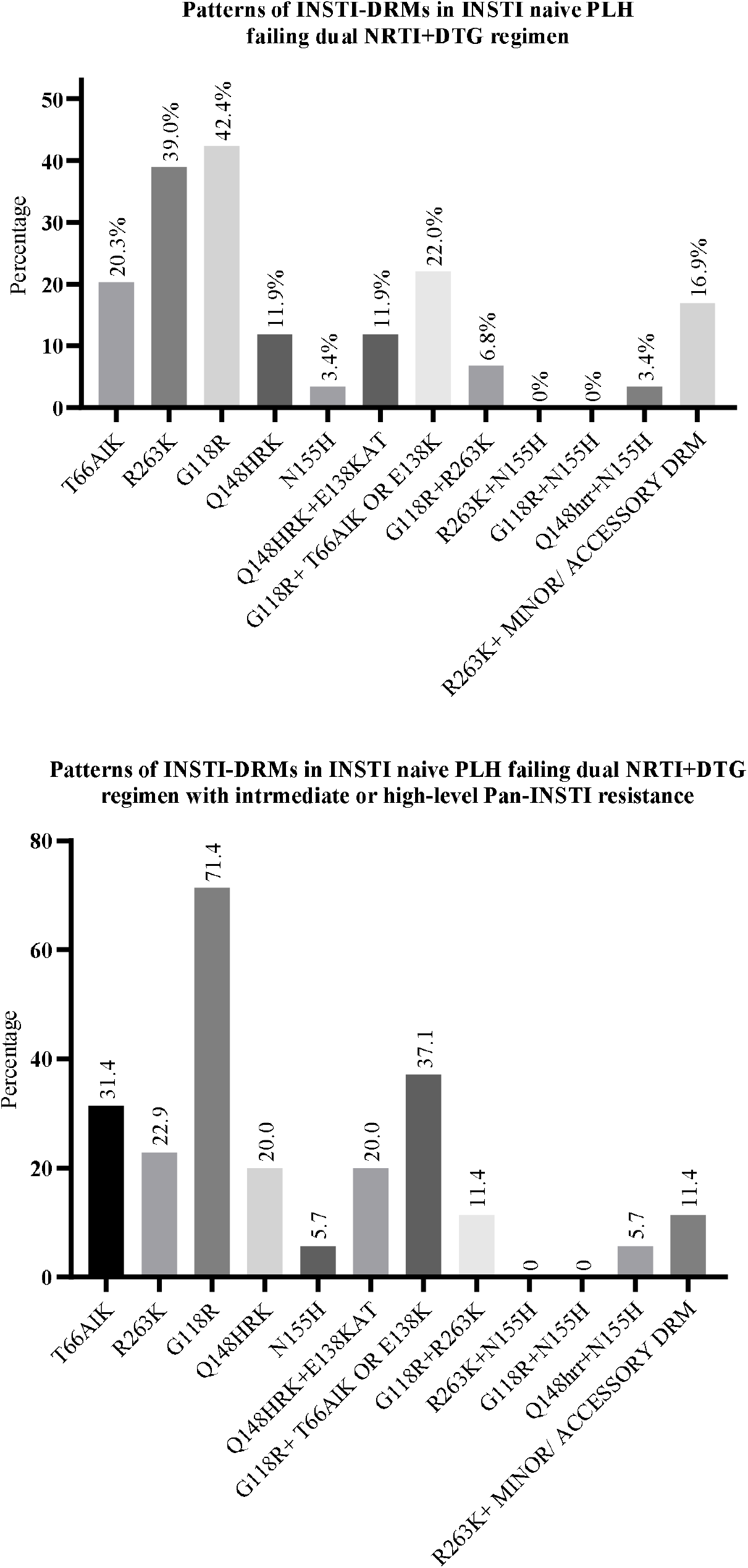
Patterns and relative frequencies of signature Integrase Strand Transfer Inhibitor (INSTI) drug resistance mutations (DRMs) in all patients and in patients with high-level pan-INSTI resistance. The top panel shows data from all patients, while the bottom panel shows data from patients with high-level pan-INSTI resistance. Additional patterns observed, but not represented in the figure, include E92EQ + G118GR (n=1), L74M + G118R (n=1), E138K + G140E + D232N (n=1), E138A + G149A (n=1) and G140R + G163R (n=1). Major INSTI mutations, as defined by the Stanford University HIV Drug Resistance Database^[14]^, include T66I, E92Q, G140S, Y143CHR, S147G, Q148HRK, and N155H. **Abbreviations**: INSTI – Integrase Strand Transfer Inhibitor; DRM – Drug Resistance Mutations; PLH – People Living with HIV; NRTI – Nucleoside Reverse Transcriptase Inhibitor; DTG – Dolutegravir.

Among specific patterns of INSTI DRMs, **R263K** occurred in combination with other accessory/minor DRMs in 16.9% of patients (n=10). These include The **R263K** DRM occurred with **M50I** (n=4), **E157Q** (n=4) and **E138K** (n=1). The **G118R+ R263K + E138K** DRMs occurred in 6.8% (n=4) patients, and **G118R+ T66AIK** or **E138K** in 22.03% of patients (n=13) [**G118R+E138K+T66I**: 18.6% (n=11), **G118R+E138K**: 5.1% (n=3)]. Isolated **G118R** DRM occurred in only five patients, compared to isolated **R263K** DRM in nine patients. The **Q148HRK** signature INSTI DRM exclusively appeared in combination with **E138KAT**, another signature INSTI DRM, in 11.9% of patients (n=7). The pattern of **N155H+R263K** and **N155H+G118R** were not observed. . Specific DRMs and patterns are described in **Figure-2** and **Supplementary Table S1**.

### Emergence of pan-INSTI resistance

Overall, 59.3% (n=35) patients who developed major INSTI DRMs on a failing ART regimen of dual NRTI + DTG regimen developed pan-INSTI resistance, defined as intermediate-level (ILR) or high-level (HLR) resistance to all first-and-second-generation INSTI agents.

Among these patients, majority (657%, n = 23), of patients developed the **G118R** DRM. Notably, **G118R + E138K** was present in 37.1% (n = 13) of patients with pan-INSTI resistance. The **G118R+R263K** DRMs occurred in 11.4% (n=4), and R263K+E157Q and R263K+E138K, both occurred in two patients. (**Figure-2 and Supplementary Table-S1**).

HIV subtype was reported for 41 patients (Subtype-B: n= 14, Non-B: n=27). It was observed that G118R and **Q148HRK** DRMs occurred more frequently in non-B subtypes (**R263K**: 37.0% [n=10], **G118R**: 48.2% [n=13], **Q148HRK**: 14.8% [n=4]) compared to a higher frequency of occurrence of **R263K** mutation in subtype-C (**R263K**: 50.0% [n=7], **G118R**: 35.7% [n=5], **Q148HRK**: 7.1% [n=1]) (**Figure-3 and Supplementary Table-S1**).

**Figure 3:**
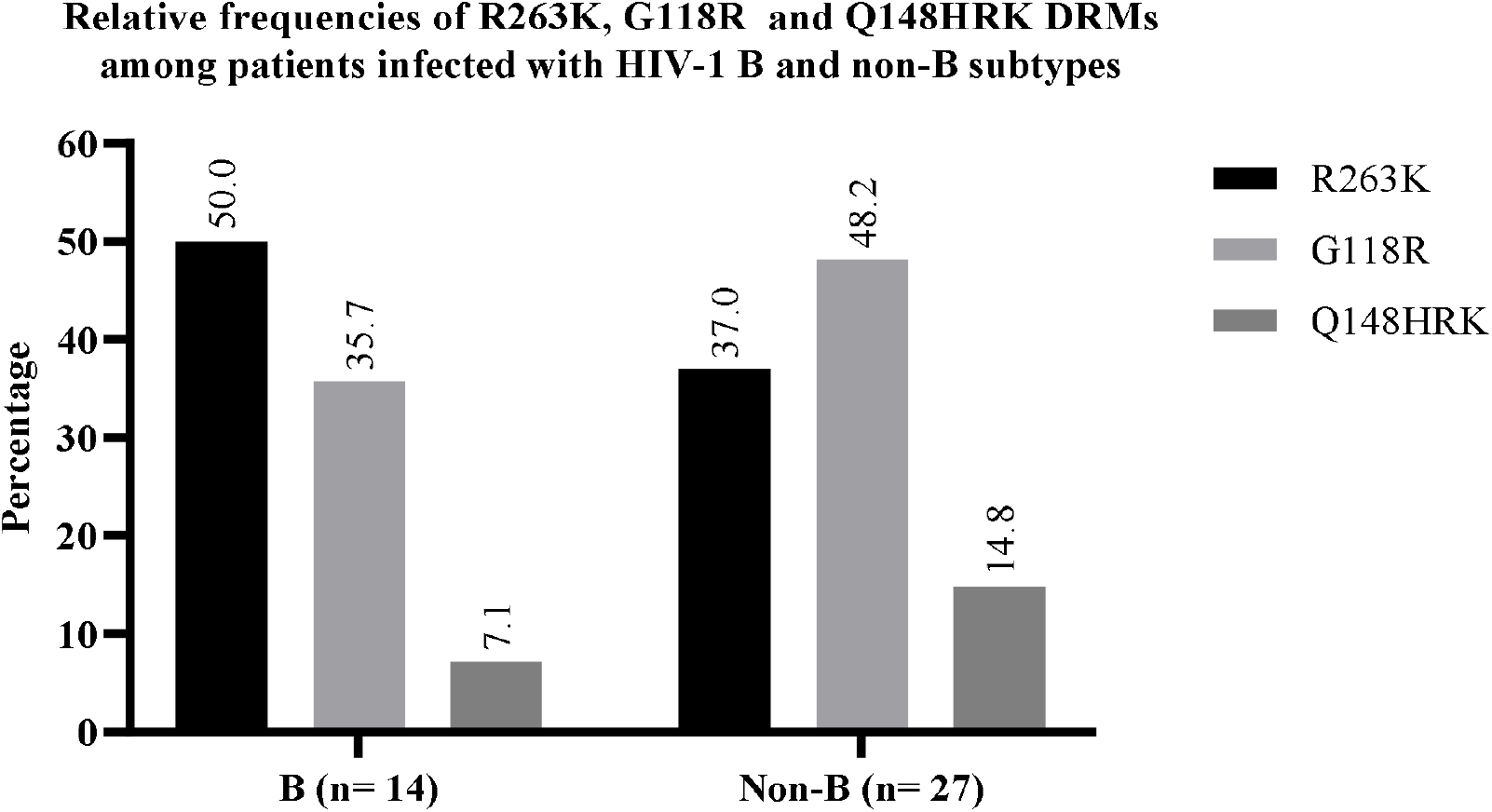
Relative frequencies of R263K, G118R, and Q148HRK DRMs in patients infected with HIV-1 subtype B compared to non-B subtypes. Non-B subtypes included A, C, F, G and circulating recombinant forms(CRF). Data from 42 patients with known HIV-1 subtype are presented, excluding one patient with a B/C recombinant strain. The bars represent the frequencies (%) of each mutation.

No difference was observed in geographic region, virological status, and NRTI backbone between those developing R263K compared with G118R.

## Discussion

This systematic review of published literature reports on INSTI-naïve PLH who experienced virological failure on the WHO recommended dual-NRTI+ DTG based ART regimens with emergent INSTI-DRMs. It describes the frequencies of occurrence of the four signature INSTI-DRMs, the patterns of their occurrence in combination with other major and minor DRMs, and the DRMs responsible for pan-INSTI resistance. We specifically studied INSTI-naïve PLH failing on a dual NRTI + DTG based ART, as the same regimen is recommended by the WHO as the preferred ART for all populations.^[11]^ This study also explores the relative frequencies of various INSTI DRMs in association with HIV-1 subtypes. Overall, this review describes 59 unique patients, reported in 21 publications across 19 studies.^[16,17,26–35,18,36,19–25]^

Previous studies have identified four signature DRMs detected in INSTI-naïve PLH failing on DTG-based ART, which include the R263K, G118R, Q148HRK, and N155H DRMs, with the first three being relatively commoner amongst previously INSTI-naive PLH.^[10,37]^ In our study as well, DRMs clustered around these four major pathways, with G118R being the most prevalent, followed by Q148HRK and R263K.

The R263K DRM has been previously described as the most common DRM developing in PLH failing a DTG-based ART, occurring in previously INSTI-naïve patients who received DTG under various scenarios, including DTG based three-drug ART, DTG+3TC dual ART, DTG monotherapy, and a DTG + optimised background therapy (OBT).^[10,38]^

The R263K DRM is often considered as a DTG-specific DRM and is selected primarily by second generation INSTIs besides being classified as a secondary EVG-selected DRM. It confers modest resistance to DTG when occurring in isolation, contributing to a median reduction in dolutegravir susceptibility of ∼2-fold.^[38,39]^ The presence of secondary DRMs concurrently with R263K represents a complex interplay between reduced susceptibility and replication fitness cost to the virus. In vitro susceptibility results revealed ∼ 5-fold reduction in susceptibility to DTG for isolates with R263K plus two additional mutations, with specific co-existing DRMs contributing differently to the activity of R263K-containing integrase^[3,5]^, particularly, M50I, which did not influence strand transfer activity.^[40]^ The combination of E138K and R263K decreased integrase strand-transfer activity to about 60% compared to the WT enzyme and failed to restore viral infectivity.^[41]^ Conversely, two mutations which co-occurred with R263K; N155H and E157Q, have previously been shown to be synergistic i.e E157Q compensates for the reduced activity of R263K-containing integrase, and N115H partially compensates for the fitness cost imposed by R263K.^[42,43]^ We observed that R263K in combination with M50I or E138K occurred in 10 out of 23 patients, with R263K+ E157Q occurring in only four patients, and R263K + N155H occurring in no INSTI-naïve PLH failing dual NRTI+ DTG-based ART.

The G118R DRM, unlike R263K, alone confers high-level resistance, and is associated with a significant reduction of HIV replication capacity.^[44,45]^ The G118R DRM has been reported previously in INSTI-naïve PLH failing a DTG-based ART regimen. In a recent systematic review including 99 INSTI-naïve PLH failing DTG based ART by Tao et al, G118R occurred nearly half as frequently as R263K, which occurs from a single nucleotide transition and represents a conserved sequence, compared to G118R, which arises from transversion and involves the substitution of an amino acid with markedly different biochemical properties compared with wildtype.^[10]^ Another systematic review by Soo-Yon Rhee et al, among INSTI-naive PLH with active virus replication receiving a standard dolutegravir (DTG)-containing regimen also observed R263K to occur more frequently than G118R.^[37]^ In our study, G118R was the most common DRM in previously INSTI-naïve PLH failing a dual NRTI + DTG based ART, which occurred marginally more frequently than R263K. This difference, compared to previous reports possibly arises from the specific focus of our study, which includes patients failing a dual NRTI + DTG based ART and excludes other regimens including dual DTG+3TC, DTG monotherapy and DTG+ OBT. This observation is relevant to the real world setting especially in low-middle income countries (LMIC), where DTG use with a dual NRTI backbone is most often undertaken without baseline drug-resistance testing, which might entail including potential suboptimal background therapy, with the acquisition of G118R DRM being dependent on genetic polymorphisms and monotherapy.^[46]^

The G118R DRM has the strongest impact on DTG susceptibility.^[7]^ It has however, conventionally been considered as rarely selected by DTG owing to its fitness cost to the virus. The relative rarity of G118R may also be due to its usual requirement for mutations at two nucleotides rather than one nucleotide, regardless of subtype.^[38,46]^ Moreover, the selection of G118R in certain settings and not others is most likely due to codon usage at position 118; although rare in certain subtypes of HIV-1, the presence of the GGA (G) codon is favourable to a transition to AGA (R).^[3]^ Moreover, HIV-1 subtypes have been postulated to affect the selection of the G118R DRM, with G118R being more often selected in non-B subtypes, possibly linked to natural polymorphisms.^[7,44–49]^ However, the evidence remains scanty, and the exact mechanism is not known. At present there remains insufficient data to determine whether a particular subtype is predisposed to the emergence of specific mutations. In our study, it was observed that the G118R DRM occurred more frequently in INSTI-naïve PLH failing a dual NRTI + DTG based ART, among patients for whom HIV-1 subtype was available.

As mentioned prior, G118R is a sinister mutation that confers pan-INSTI resistance. When the G118R mutation occurs concurrently with the T66IA and E138K mutations, this results in a 6.5 to 22-fold resistance to DTG. In our study, it was also observed that G118R commonly existed concurrently with the E138K and T66I DRMs, which has been shown to partially restore the replicative fitness of G118R-containing integrase.^[31,50]^ Further characterisation of the effect of various secondary DRMs on the replicative fitness of HIV with the G118R mutation is required, since the effect of G1118R on viral fitness may also be subtype dependent.^[45,51]^ These findings indicate the need for close monitoring of dolutegravir implementation in the region, as there is still inadequate information on INSTI-resistance mutations that may lead to treatment failure in patients infected with non-subtype B viruses.

Our systematic review adds to the works by Tao, at al^[10]^ and Rhee, et al^[37]^ by reviewing the most commonly occurring DRMs after failure of a dual NRTI+ DTG-containing ART regimen in previously INSTI-naïve PLH and provides a real world perspective. Our review also analyses the patterns of DRMs occurring in such patients and provides preliminary evidence of variations in these patterns by HIV-1 subtype. The increasing trend in occurrence of the G118R DRM especially requires more vigil, especially in LMICs.

The limitation of our study includes the exclusion of data on the total number of patients who received the study regimen of dual NRTI+ DTG. DTG resistance has been majorly reported in uncontrolled trials, observational studies, and case reports, making collection of such data difficult. Our search identified only those studies that contained cases of emergent DTG resistance and did not identify studies in which DTG was received, but INSTI-associated DRMs did not arise. For these two reasons, a different systematic review would be required to determine the prevalence of emergent DTG resistance in different clinical scenarios. Another limitation of our study is the slight underrepresentation of HIV-1 subtype-B and the non-availability of HIV subtype in certain patients.

## Conclusion

In conclusion, this review highlights the emergence of INSTI DRMs in HIV-1 patients receiving dual NRTI + dolutegravir (DTG)-based ART, with G118R, R263K, E138K, and T66I, being commonly identified, with patterns varying by HIV-1 subtype. These findings emphasize the need for ongoing monitoring of INSTI resistance, particularly in low-resource settings where DTG is commonly used without baseline resistance testing. While the study has limitations, including underrepresentation of certain HIV subtypes, it provides valuable real-world insights to guide clinical management of drug-resistant HIV.

## Supporting information

Study Methodology Proposal

## Data Availability

Data will be made available upon reasonable request.

## Ethical Disclosures

The study comprises retrospective analysis of published deidentified data, therefore consent and IEC was waived. Study details available at OSF for further perusal.

## Declaration of generative AI and AI-assisted technologies in the writing process

During the preparation of this work the author(s) used Generative Pre-trained Transformer-3.5 [GPT-3.5, OpenAI (2023)], which was used solely for language guidance and correction. The authors confirm that no exclusively AI-generated text or figures have been incorporated into any part of the manuscript. After using this tool/service, the author(s) reviewed and edited the content as needed and take(s) full responsibility for the content of the publication.

