## Supplementary material for "Worrisome Emergence of Pan-INSTI Resistance: A Systematic Scoping Review of Dolutegravir Resistance in INSTI-Naïve Patients Post-Therapy Failure": Study Methodology Proposal

**Scoping Review Proposal: Pan-INSTI Drug Resistance in HIV Patients Following Virological Failure on Dolutegravir-Based ART**

**Abstract**
This protocol outlines a systematic scoping review of the emergence and patterns of pan-integrase strand transfer inhibitor (INSTI) resistance in HIV-1 patients on Dolutegravir (DTG)-based ART. Given clinical concerns about resistance rates far exceeding those reported in clinical trials, this review will synthesize evidence from clinical trials, observational studies, case reports and relevant grey literature published to date.

**1.	Background**

**1.1	Rationale**

Dolutegravir (DTG), a widely used second-generation INSTI, has significantly improved HIV management due to its high genetic barrier to resistance, efficacy, and tolerability. However, real-world data have shown that pan-INSTI resistance can emerge at rates unexpectedly higher than those observed in clinical trials. Understanding the prevalence and patterns of drug-resistance mutations (DRMs) is crucial, especially for settings where DTG is the cornerstone of ART.

**1.2 Objective**

To systematically review and characterize the rates and patterns of pan-INSTI resistance mutations in HIV patients receiving DTG-based ART untill 31 October 2024.

**1.3 Review Question**

What are the rates and specific mutations associated with pan-INSTI resistance in HIV patients on DTG-based ART, and how do these compare with resistance data from clinical trials?

**2. Methods**

**2.1 Protocol and Registration**

The review will follow the Preferred Reporting Items for Systematic Reviews and Meta-Analyses (PRISMA) guidelines and will be registered.

**2.2 Search Strategy**

Searches will be conducted in PubMed and Google Scholar using a combination of controlled vocabulary (MeSH terms) and free-text terms. The search will target literature till 31 October 2024, with the following terms:

- **PUBMED Search Terms**:
  ("Dolutegravir"[Title/Abstract] OR "HIV Integrase Inhibitors"[MeSH Terms] OR "INSTI"[Title/Abstract] OR "Integrase Inhibitors"[Title/Abstract])
  AND ("Drug Resistance, Viral"[MeSH Terms] OR "HIV Drug Resistance"[Title/Abstract] OR "Drug Resistance"[MeSH Terms] OR "Drug Resistance Mutation*"[Title/Abstract] OR "HIVDR"[Title/Abstract] OR "Resistance Mutations"[Title/Abstract] OR "Mutation"[MeSH Terms] OR “Dolutegravir Resistance” [Title/Abstract] OR “Resistance” [Title/Abstract])
  AND ("HIV"[MeSH Terms] OR "HIV"[Title/Abstract] OR "HIV-1"[MeSH Terms] OR "HIV Infections"[MeSH Terms] OR "Anti-HIV Agents"[MeSH Terms])
  AND ("case reports"[Publication Type] OR "clinical study"[Publication Type] OR "clinical trial"[Publication Type] OR "observational study"[Publication Type] OR "multicenter study"[Publication Type] OR "comparative study"[Publication Type])
  NOT ("systematic review"[Publication Type] OR "meta analysis"[Publication Type] OR "review"[Publication Type])
  AND ("2019"[Date - Publication] : "2024"[Date - Publication])
- **GOOGLE SCHOLAR Search Terms**:
  ("Dolutegravir" OR "Integrase strand transfer inhibitor" OR "HIV Integrase Inhibitors" OR "INSTI" OR "Integrase Inhibitors")
  AND ("Drug Resistance, Viral" OR "HIV Drug Resistance" OR "Drug Resistance Mutation*" OR "HIVDR" OR "Resistance Mutations" OR "Mutation")
  AND ("HIV" OR "HIV-1" OR "HIV Infections" OR "Anti-HIV Agents")
  AND ("case reports" OR "clinical study" OR "clinical trial" OR "randomized controlled trial" OR "observational study" OR "multicenter study" OR "comparative study")
  NOT ("systematic review" OR "meta analysis" OR "review")

**2.3** **Eligibility Criteria**

- **Inclusion:** Case reports, observational studies, and trials reporting on pan-INSTI resistance mutations in HIV-1 patients receiving DTG-based ART, including relevant grey literature such as conference papers and abstracts.
- **Exclusion:** Reviews, meta-analyses, studies without detailed resistance mutation data, studies not reporting individual patient data, or lacking essential data such as ART history and viral load information. Studies not involving patients treated with DTG were excluded. When HIV-1 isolates with INSTI-associated DRMs appeared in multiple publications from the same clinical trial or cohort, the isolates were linked to the first publication.

**2.4 Data Extraction and Management**

**2.4.1 Data Extraction Process**

The data extraction will follow the CoCoPop framework:

- **Population:** HIV-1 infected individuals on dual NRTI+DTG-based ART for first- or second-line treatment, geographic region, age, gender, and viral load.
- **Condition:** Presence of pan-INSTI resistance mutations, associated clinical outcomes, weeks (24, 48, 96, and so on) after initiation of DTG when virological failure occurred, author-defined virological failure criteria, type of drug resistance testing (DRT) performed, and emergent INSTI DRMs. For people living with HIV (PLWH) who had more than one isolate with INSTI-associated DRMs, a cumulative count of DRMs from all isolates was considered, provided the isolates were from the same period of virological failure on the study regimen. For this analysis, mutations present as part of a mixture with wildtype were considered mutant.
- **Context:** ART regimens and duration, prior exposure to other INSTIs, relevant demographic data, co-administered Rifampicin, and adherence to ART

**2.4.2 Data Extraction Form**

A standardized data extraction form will be utilized, capturing the following:

•	Study Identification: Title, authors, year, publication source, and study design

•	Patient Demographics: Age, gender, viral load, and ART history

•	Resistance Mutations: Specific mutations, classification (major/minor), and impact on treatment

•	Study Methods and Quality: Methodological details, potential sources of bias, and study limitations

**2.4.3 Reviewer Process**

Two reviewers will independently extract data. Discrepancies will be resolved by discussion, with a third reviewer (expert in the field) consulted if necessary.

**3. Study Methods, Data Quality, and Potential Sources of Bias**

| **Study Type** | **Potential Bias** | **Bias Mitigation** |
| --- | --- | --- |
| **Case Reports/Series** | Selection, Reporting | Prioritize detailed reports; acknowledge potential reporting bias |
| **Observational Studies** | Selection, Information | Prioritize well-conducted studies; ensure clear definitions |
| **Clinical Trials** | Selection, Attrition | Scrutinize eligibility criteria; address attrition bias |
| **Errata and Letters** | Information | Evaluate completeness of information |
| **Conference Abstracts** | Selection, Information | Consider limitations in data interpretation |
| **Grey Literature** | Publication | Assess relevance and acknowledge publication bias |

**4. Risk of Bias Assessment**

Qualitative synthesis

**5. Data Synthesis Strategy**

Data will be synthesized using descriptive and qualitative methods, with subgroup analyses based on study design and geographic setting.

**6. Analysis Plan**

**6.1 Effect Measures:** Descriptive analysis focusing on mutation frequency and patterns

**6.2 Sensitivity Analysis:** NA

**6.3 Subgroup Analysis:** Compare mutation patterns across different settings and patient populations

**7. Reporting**

The review will adhere to PRISMA guidelines, with findings presented in tables and figures highlighting resistance mutations and prevalence.

**8. Timeline**

•	Protocol development: 1 month

•	Literature search: 2 months

•	Data extraction: 3 month

•	Data synthesis: 2 month

•	Manuscript preparation: 1 month

•	Total: 9 months

**9. Ethics and Dissemination**

No ethical approval needed as the review uses publicly available data. Results will be disseminated through a peer-reviewed journal and presented at conferences. Ethics compliance of individual studies will be scrutinized.
